# High-resolution ultrasonography of gingival biomarkers for periodontal diagnosis in healthy and diseased subjects

**DOI:** 10.1101/2021.11.08.21265833

**Authors:** Colman Moore, Jane Law, Christopher Pham, Kai Chiao Joe Chang, Casey Chen, Jesse V. Jokerst

## Abstract

Periodontal disease affects nearly 50% of Americans but diagnostic methods have remained the same for decades. Periodontal examination via physical probing provides critical metrics such as pocket depth, clinical attachment level, and gingival recession; however, this practice is time consuming, variable, and often painful. In this study, we investigated high-frequency ultrasound (40 MHz) for the image-based measurement of periodontal metrics. Imaging was performed at midbuccal sites for a set of periodontally healthy (n = 10) and diseased (n = 6) subjects and image-based measurements were compared to gold-standard physical probing measurements. Human operators identified relevant markers (e.g., cementoenamel junction, gingival margin, alveolar bone crest) in B-mode ultrasound images from 66 teeth to calculate gingival height and alveolar bone level. These metrics were correlated to clinical measurements of probing pocket depth and clinical attachment level for disease staging (1.57-mm bias and 0.25-mm bias, respectively). Interoperator bias was negligible (<0.1 mm) for gingival height measurements and 0.45 mm for alveolar bone level measurements. The ultrasonographic measurements of gingival height and alveolar bone level served as effective diagnostic surrogates for clinical probing measurements while offering more detailed anatomical information and painless operation.

## INTRODUCTION

Nearly 50% of Americans have periodontitis^1^ resulting in pain, tooth loss^2, 3^, reduced quality of life^4-7^ and even systemic effects like cardiovascular disease^8^, but tools to diagnose/monitor periodontitis have major limitations. Clinical assessment (by periodontal examination) and radiography are currently the standard of care but suffer from being time-consuming for the clinician, uncomfortable for the patient, and subject to large errors—inter-operator variation in probing can be >40% ^9^. Moreover, clinical assessment and radiographic examination may not capture all clinical information (e.g., gingival thickness and inflammation).

The periodontal examination provides critical information such as probing pocket depth (PPD; current periodontal health) and clinical attachment level (CAL; cumulative destruction)^10^. PPD, CAL, and other clinical parameters form the basis of periodontal diagnosis. Radiography offers excellent sensitivity to hard tissue (bone, enamel, etc.) but cannot discriminate between healthy and diseased gingiva or map disease within soft tissue; it also has a small but non-negligible dose of ionizing radiation.

The use of ultrasound imaging to resolve dental and periodontal structures, especially for alveolar bone and the cementoenamel junction, has been demonstrated in recent years ^11-18^. It has also been used for imaging peri-implantitis and the topography of edentulous crestal bone ^19-21^. Ultrasound imaging has the benefits of being a portable and low-cost alternative to radiography that is noninvasive and free of ionizing radiation, and can measure the thickness of the gingiva, which is deemed difficult with radiographic imaging modalities. We and others have previously evaluated these techniques via small animal models and case studies but higher powered studies in human subjects are relatively rare ^22-24^. Here, our goal was to evaluate the value of high frequency ultrasound in periodontology in a clinical context. We achieved this goal by identifying and evaluating imaging biomarkers and comparing these biomarkers to established clinical metrics of periodontal health.

## METHODS

### Materials

A high-frequency, commercially available imaging ultrasound system was employed (Vevo LAZR, Visualsonics, Toronto CA) with a 40-MHz linear array transducer (LZ-550). Disposable tegaderm films were used as sterile transducer sleeves (3M, Minnesota, USA). Periodontal probing measurements were conducted with a Williams and Marquis probe. Extracted swine jaws were provided by Sierra For Medical Science, Inc. Whittier, CA.

### Subject recruitment and clinical examination

The study protocol received approval from the USC and UCSD Institutional Review Boards and was in accordance with the ethical guidelines for human subjects research established by the Helsinki Declaration of 1975. The study subjects were identified from patients seeking dental care at the Herman Ostrow School of Dentistry. As part of the clinical protocol, the patients routinely received comprehensive extra- and intra-oral examinations, medical and dental history review, a set of full-mouth radiographs, periodontal examination, periodontal diagnosis, and treatment planning. Eligible subjects were healthy adults who weighed at least 110 pounds with one quadrant with at least upper and lower anterior teeth. Subjects were excluded if they had bloodborne pathogen infections, bleeding disorders, acute oral infections, or were pregnant or lactating women. Two subject groups were recruited based on the periodontal diagnosis described in the 2017 World Workshop on the Classification of Periodontal and Peri-implant Diseases and Conditions^25^. The first group (n = 10) comprised subjects with the following diagnosis: periodontal health in intact or reduced periodontium in stable periodontitis patients, or dental biofilm-induced gingivitis in the intact or reduced periodontium. The second group (n = 6) comprised subjects diagnosed with periodontitis, Stage II-IV, and Grade B or C, with localized or generalized involvement.

The periodontal diagnosis was reviewed and confirmed by the examiners. The maxillary or mandibular six anterior teeth were then selected for the study. Periodontal probing depth was determined with a Williams and Marquis probe at six sites per tooth (mesio-labial, mid-labial, disto-labial, mesio-lingual, mid-lingual, and disto-lingual). Tooth mobility was determined by Class 1: mobility of up to 1 mm in an axial direction, Class 2: mobility of greater than 1 mm in an axial direction, and Class 3: mobility in an apico-coronal direction (depressible tooth). The bleeding on probing (BOP) provoked by applying a probe to the bottom of a sulcus/pocket was recorded. Gingival recession was recorded by measuring the distance between the CEJ to the top of the gingival margin in the mid-labial aspect of the tooth with a periodontal probe. Clinical attachment level (CAL) was determined from CEJ to the bottom of the pocket. The gingival phenotype was determined by inserting the periodontal probe into the mid-labial surface of the tooth. A thin gingival phenotype was assigned if the probe was visible through the gingival tissue.

### Periodontal ultrasound imaging

Subjects were seated in the supine position in the dental chair and imaged with a handheld, linear array transducer by a clinician. A disposable transparent sleeve was used to wrap the transducer in addition to sterile ultrasound coupling gel. Imaging was performed manually by positioning the transducer parallel to the long axis of the tooth along the buccal midline. A layer of ultrasound coupling gel approximately 5 mm thick was placed between the contact areas and the transducer to achieve good coupling and optimum resolution. The freehand scanning of each tooth was exported as a video file consisting of at least 1,000 frames.

### Image analysis

All imaging measurements were performed in duplicate by two individual, blinded analysts to assess human variation in the identification of anatomical biomarkers. The first was a clinician with no ultrasound experience (Analyst 1) while the second was an ultrasound researcher with no clinical experience (Analyst 2). Imaging measurements were performed digitally in the VisualSonics software and ImageJ. The distance from the gingival margin (GM) to the alveolar bone crest (ABC) was defined as the image-based gingival height (iGH). Similarly, the distance from the CEJ to the ABC was defined as the image-based alveolar bone level (iABL). The image-based gingival thickness (iGT) was measured at the midpoint of the ABC and GM. All images had to meet specific quality criteria prior to measurement. These were: 1) clear resolution of the GM, (2) clear resolution of the ABC, and (3) a lack of interfering artifacts coincident with the relevant anatomy. If these conditions were met, then imaging measurements were performed (**Table S1**).

## Results AND DISCUSSION

This study enrolled 16 random subjects for periodontal ultrasound imaging during dental care appointments at a clinic. The subjects were classified as healthy (n = 10) or diseased (n = 6) following oral examination as described in the methods. It is worth noting that the disease severity (i.e., PPD, CAL, bone loss) of the teeth may not be consistent with the diagnosis, which was derived from assessment of the whole mouth. A high-frequency, commercially available imaging system was used for chairside imaging of subjects (**Fig. 1A**). The handheld linear array transducer (40 MHz, **Fig. 1B**) permitted access to the maxillary/mandibular incisors and cuspids (teeth 6-11 and 22-27, **Fig. 1C**). B-mode images (2D US cross sectional images) were collected in the sagittal plane at the midbuccal site of each tooth. The anatomy of the imaged region is depicted in **Fig. 1D** for comparison to a representative B-mode image in **Fig. 1E**. In general, six anatomical markers were consistently identified and used to orient the imaging operator/analyst: the alveolar bone, the gingiva, ABC, GM, CEJ, and the tooth surface (**Fig. 1D-E**).

**Figure 1.**
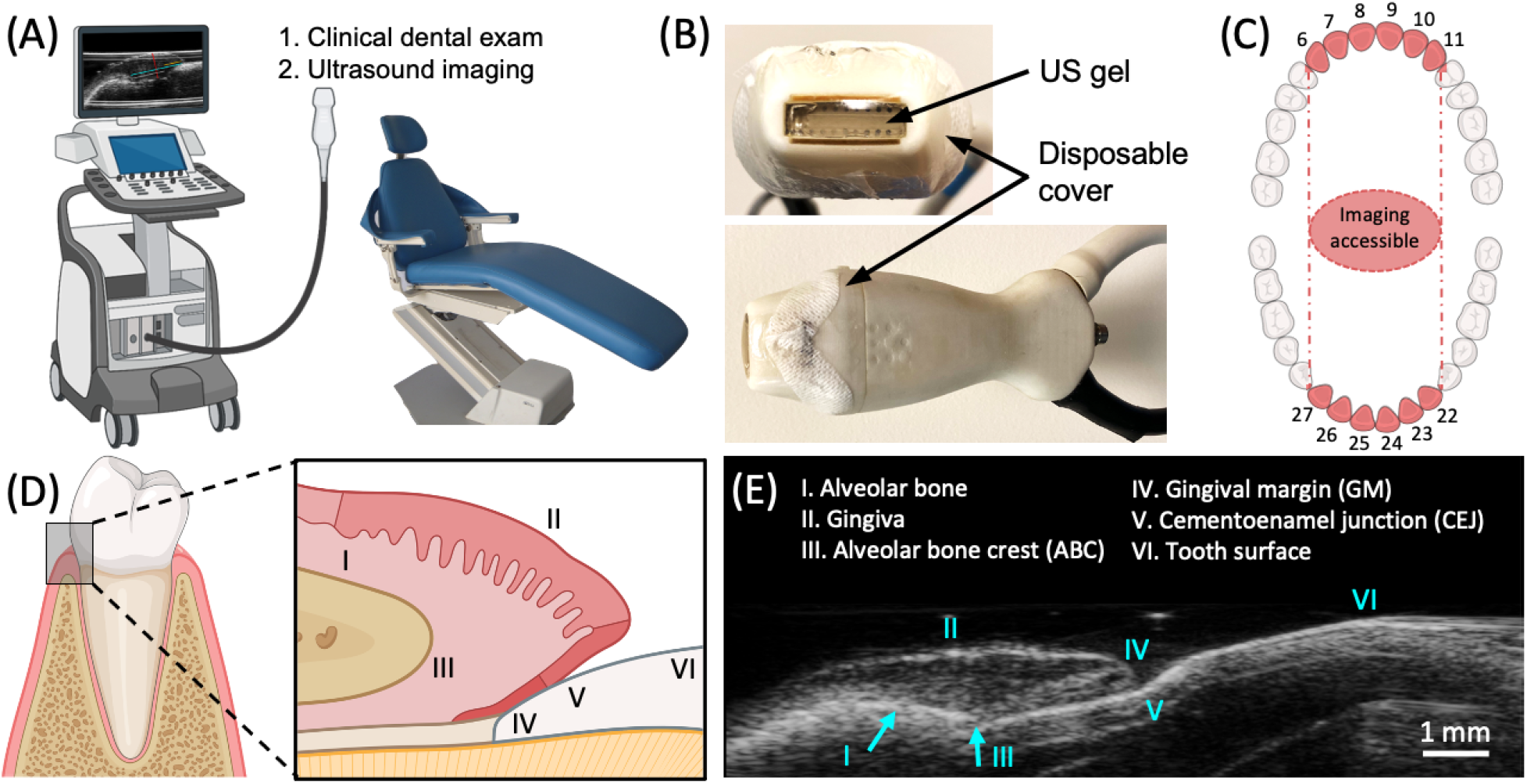
Overview of periodontal ultrasound imaging. (**A**) Schematic of chairside ultrasound imaging during routine dental examination. (**B**) Photograph of the commercial 40-MHz transducer with coupling gel and sterile sleeve used for subject imaging. (**B**) Dental chart with teeth highlighted (6-11, 22-27) that could be physically accessed by the US transducer. (**C**) Diagram of the periodontal anatomy surrounding the gingival sulcus with magnification of the sagittal plane. Roman numerals denote the I: alveolar bone, II: gingiva, III: alveolar bone crest (ABC), IV: gingival margin (GM), V: cementoenamel (CEJ), VI: tooth surface. (**D**) B-mode ultrasound image of the region in (C) for the central mandibular incisor (#25) of a patient with anatomical markers labeled.

In this work, the identification of the CEJ in relation to the ABC and GM was of particular interest. Locating the CEJ is important for determining metrics of periodontal health such as gingival recession and clinical attachment level (CAL). Since the CEJ is typically covered by the gingiva (subgingival), its exact location is difficult to determine via physical probing and can be subject to significant error—for midbuccal sites, Vandana et al. reported over- or underestimation of the CEJ by trained periodontists for 74% (34/46) of measured teeth^26^. However, recent reports have conclusively demonstrated that US can accurately identify the CEJ ^17, 18^. Here, we extended this concept by using 40-MHz US to locate the CEJ in relation to other anatomical markers for the image-based calculation of periodontal metrics that typically require estimation by physical probing. First, we confirmed that our ultrasound system could resolve the CEJ in extracted swine jaws (**Fig. S1**). This was achieved by imaging each tooth (**Fig. S1A**), measuring the CEJ with image analysis (**Fig. S1B-D**), and comparing the values to tactile probing measurements following gum flap resection (**Fig. S1F-G**) and measurement by a clinician (**Fig. S1H**). These measurements are restricted to integers but represent the gold standard and showed good agreement with imaging (<1.0 mm difference between GM-CEJ values and <0.5 mm difference between GM-ABC values) (**Fig. S1I**).

In humans, 79 B-mode images were acquired from 16 subjects comprising 43 teeth clinically diagnosed as healthy and 36 diagnosed with periodontal disease via physical measurements and examination. Of these images, 66 (84%) met quality criteria and were used for analysis. All image quality metrics, image measurements, and clinical measurements are included in **Table S1**.

One simple measurement is the distance between the CEJ and GM—since the CEJ is a static landmark, this value can be used to track gingival migration (recession or overgrowth). **Fig. 2** demonstrates varying positions of the CEJ relative to the GM for six different human subjects. The CEJ presents as an angled disruption in the echogenicity of the tooth surface between the GM and ABC. For subjects in **Fig. 2A-C**, the CEJ is apical to the GM (typically, a positive health marker). While a clinician will also diagnose these cases as non-recessed, it is unlikely that physical probing can locate the subgingival CEJ with the same precision and accuracy ^26^. With imaging, the exact amount of gingival overlap can be measured, providing insight into future risk. For example, **Fig, 2C** shows a tooth with only 0.6 mm of gingiva above the CEJ, while in **Fig. 2D**, the CEJ and GM are coincident. Lastly, two cases of gingival recession are presented in **Fig. 2E-F**. Here, the CEJ is coronal to the GM. Overall, these cases show the potential value of monitoring the GM to CEJ distance with US imaging for evaluating gingival recession and its future risk.

**Figure 2.**
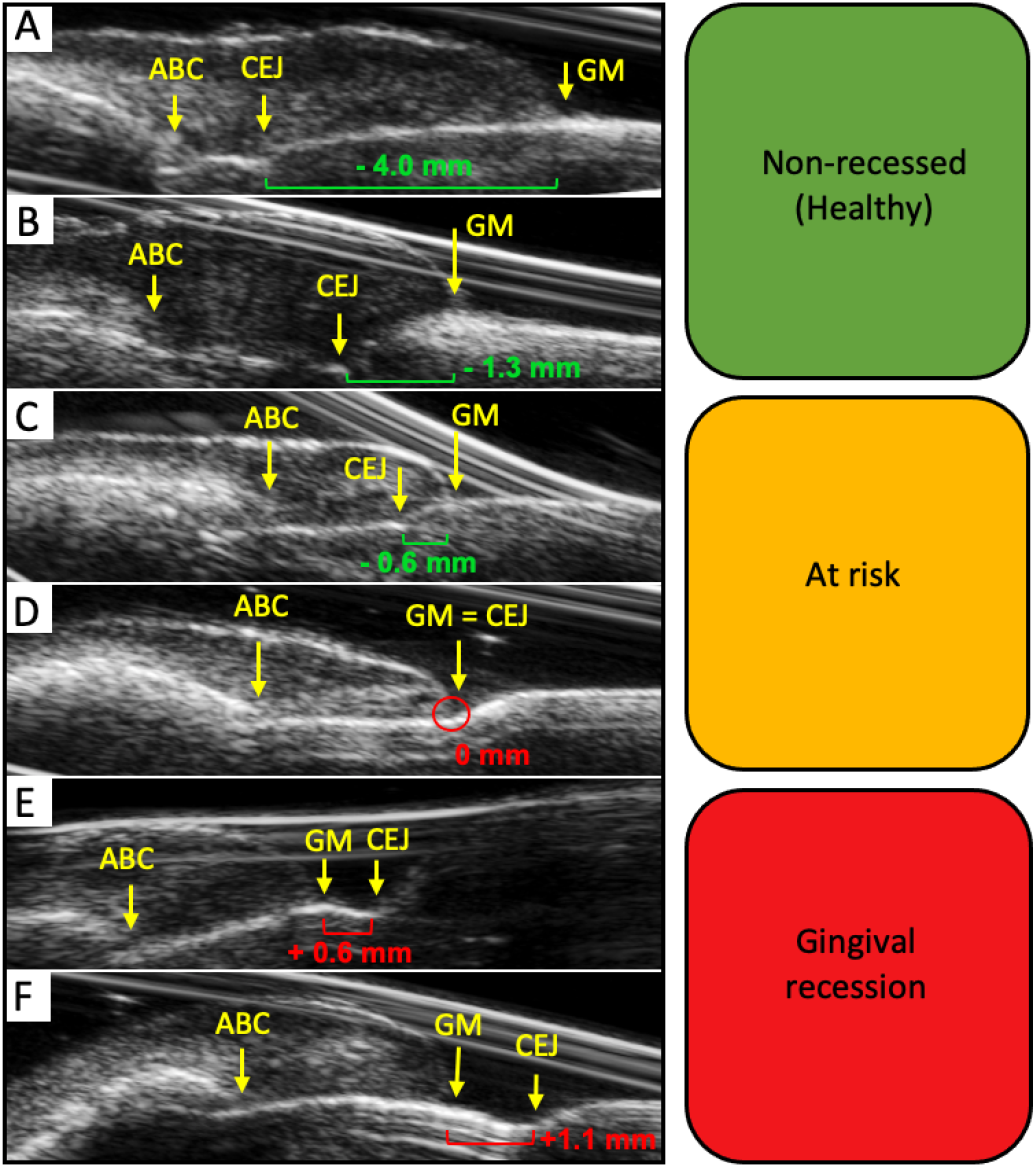
B-mode images in six subjects demonstrating US monitoring of gingival recession via periodontal landmarks. Clinically, the distance between the CEJ and GM defines the extent of gingival recession and is used to determine CAL. Panels **A-F** show teeth from subjects with increasing levels of gingival recession. (**A-C**) Images from subjects with the CEJ apical to the GM (i.e., non-recessed). These measurements are represented as negative values (green). (**D**) Image from a subject where the GM is coincident with the CEJ (i.e., PPD = CAL). (**E-F**) Images from subjects with the CEJ coronal to the GM (i.e., recessed). Recessed measurements are represented as positive values (red).

Though useful for longitudinal monitoring, the position of the CEJ in relation to the GM at a single point in time is insufficient for diagnosing periodontal health. Therefore, we evaluated the diagnostic value of measurements derived from the ABC, CEJ, and GM. Specifically, we measured the image-based alveolar bone level (iABL) and image-based gingival height (iGH) for each collected B-mode image and compared them to clinically measured PPD and CAL values. We also compared the magnitudes of these values after binning the patient images into “healthy” or “diseased” groups corresponding to the clinical diagnosis of the patient. It should first be emphasized that the periodontal pocket (or gingival sulcus) does not generate sufficient endogenous contrast to be resolved with US alone. Despite this, we hypothesized that the distance from the GM to the ABC (iGH, easily resolvable with US), could function as a surrogate measurement of the pocket depth. All image-based measurements, including iGH, iABL, iGR (image-based gingival recession), and iGT (image-based gingival thickess) are depicted in **Fig. 3A**. To determine the iGH and iABL values in this study, two blinded analysts (one clinician and one researcher) independently measured each frame, and their values were averaged. Bias between raters was < 0.1 mm for iGH (**Fig. 3B**) and 0.45 mm for iABL (**Fig. 3C**). The increased variance for iABL reflected differences between the raters in assigning the CEJ, which may be a less obvious feature than the ABC or GM.

**Figure 3.**
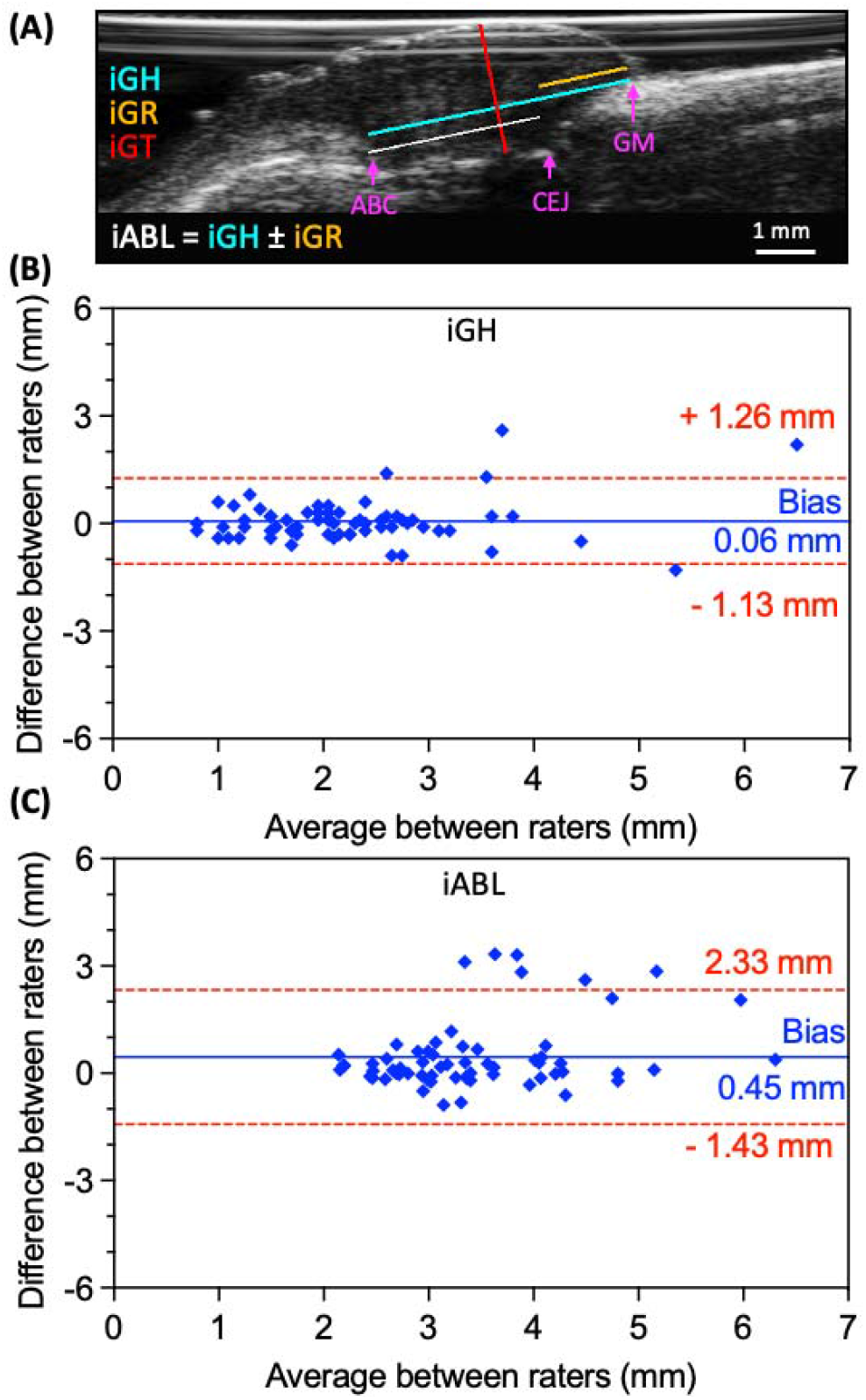
Ultrasound for diagnostic measurements and interrater variability for US image-based measurements of pocket depth (iPD) and clinical attachment level (iCAL). (**A**) B-mode image with manual annotations showing the extraction of iGH (teal), iABL (white), iGR (yellow), and iGT (red) for a representative subject. The iGH is measured from the ABC to the GM. (**B-C**) Bland-Altman plots comparing the iGH and iABL measurements from two blinded image analysts for the same image set (n = 66 teeth). The increased variance (bias) between analysts for iABL (0.45 ± 0.96 mm) relative to iGH (0.06 ± 0.61 mm) was due to differences in identification of the CEJ between the two image analysts.

The average iGH and iABL values for all teeth were compared to clinical PPD and CAL measurements, respectively, obtained by a clinician (**Fig. 4, Table S1**). The average PPD measurements were 1.68 mm for healthy subjects and 2.25 mm for diseased subjects (**Fig. 4A**). A similar increase was observed for iGH measurements: 3.19 mm for healthy subjects and 3.67 mm for diseased subjects (**Fig. 4A**). Expectedly, iGH values were larger than PD values (1.57 mm Bland-Altman bias, on average), because the iGH measurements terminated at the ABC rather than the gingival sulcus (**Fig. 4B**). Most importantly, both PD/CAL and iGH/iABL were larger for diseased than healthy patients. The CAL measurements were 1.68 mm for healthy subjects and 2.56 mm for diseased subjects (**Fig. 4C**). For iABL, the healthy average was 1.80 mm and the diseased average was 2.74 mm—this difference between groups was even more significant than the CAL measurements (**Fig. 4C**). Bland-Altman analysis revealed a minor 0.25-mm magnitude bias toward the iABL measurements (**Fig. 4D**).

**Figure 4.**
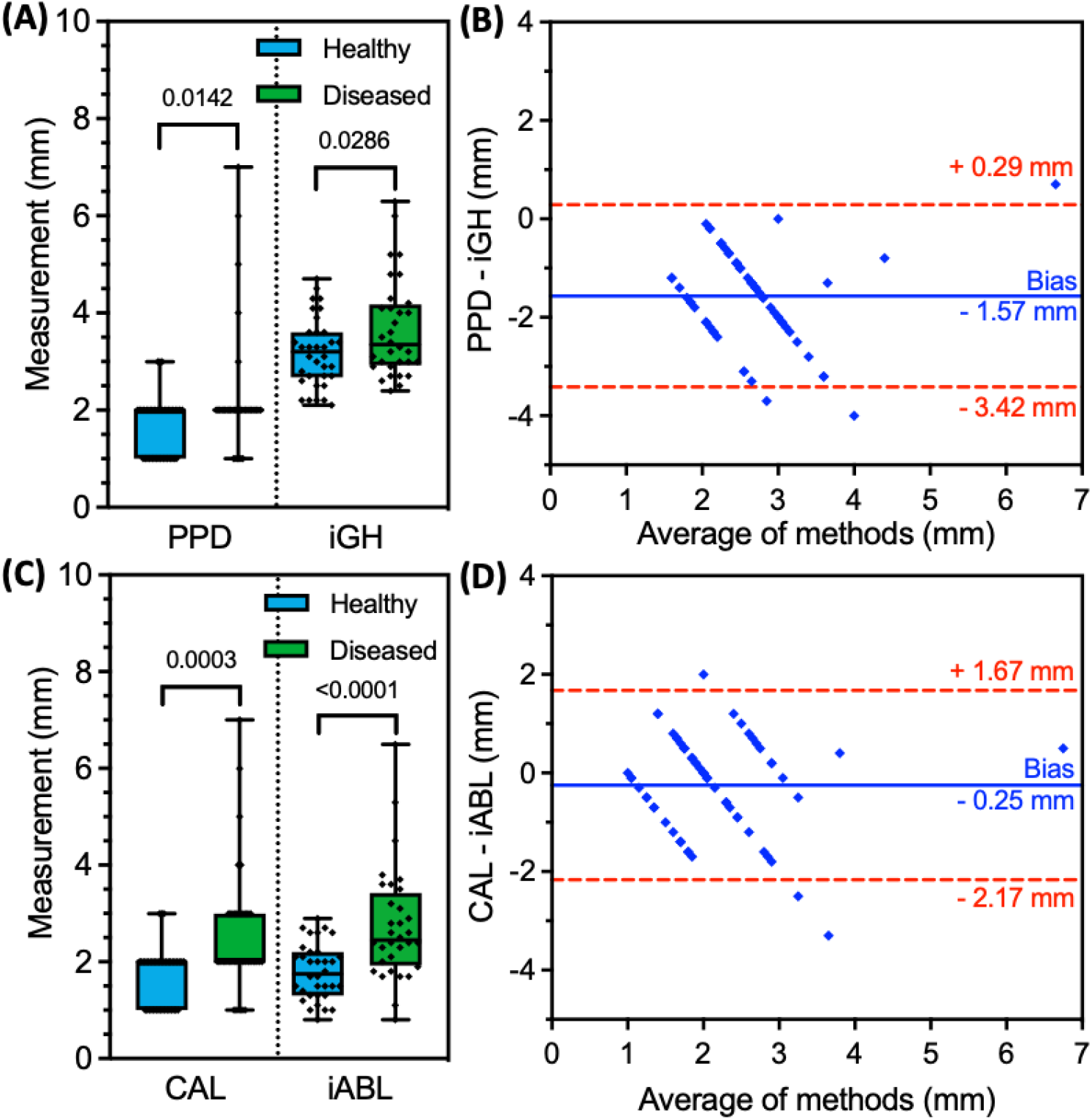
Comparison between US image-based measurements (iGH, iABL) and clinical probing measurements (PPD, CAL) for individual teeth (n = 66) of patients with healthy or diseased clinical diagnoses. (**A**) Box-and-whisker plots for PPD and iGH both indicate significantly higher measurements in the diseased group (n = 32) than the healthy group (n = 34). PPD values are limited to integers. Pairwise comparison values are p-values (unpaired t-test). (**B**) Bland-Altman analysis between the measurement methods reveal a 1.57 ± 0.95 mm bias toward iGH measurements averaged from all teeth– these values are larger because though both measurements begin at the GM, the iGH is measured to th ABC rather than the terminus of the gingival sulcus. This difference, due to the connective tissue and junctional epithelium between the ABC and gingival sulcus, is referred to as the biological width. (**C**) Box-and-whisker plots for iABL and CAL indicate significantly higher values for teeth in the diseased group than the healthy group. CAL values are limited to integers. Pairwise comparison values are p-values (unpaired t-test). (**D**) Bland-Altman analysis between the iABL/CAL measurement methods reveal a 0.25 ± 0.98 mm bias toward the iABL measurements, indicating a minimal difference between the two methods.

The rationale for comparing both iGH to PD and iABL to CAL is that these measurements are physically equivalent except for their termini— that is, iGH and iABL terminate at the ABC while PPD and CAL terminate at the bottom of the gingival sulcus. This difference, i.e., the distance between the ABC and the terminus of the gingival sulcus (corresponding to connective tissue and junctional epithelium) has been described as the biologic width ^27^. Therefore, if known, the biologic width should be subtracted as a correction factor to calculate an image-based pocket depth (iPD) from the iGH as well as an image-based clinical attachment level (iCAL) from the iABL. Thus, iPD should approximate PPD and correlate with iGH.

From our dataset, the average difference between iGH and PPD measurements was 1.57 mm (**Fig. 4B**). If this value is defined as the average biologic width and subtracted from each iGH measurement, then we obtain a set of iPD values after rounding to the nearest integer similar to rounding done when measuring the PPD. Likewise, we obtain a set of iCAL values after performing the same subtraction from the iABL data. Upon this analysis, we achieved 83% agreement between iPD and PPD values, and 49% agreement between iCAL and CAL values, where agreement was defined as ≤ 1 mm difference between paired measurements.

While our measured estimate for the biologic width falls within the range of mean values reported in a systematic meta-analysis (between 1.15-3.95 mm), disease state, tooth type, probing depth, and attachment loss can all affect the biological width^28^. The combination of these variables and the lack of precision in locating the subgingival CEJ with a periodontal probe are the likely reasons for the relatively low agreement between iCAL and CAL values. Indeed, this limitation of physical probing for CEJ identification reduces the value of this comparison. Given the higher accuracy of physical probing for PPD measurements, the agreement between iPD and PPD is both more reliable and promising.

Importantly, while recapitulating the PPD values via imaging has value, the iGH is simpler and more straightforward to measure. It also does not require *a priori* knowledge of the biological width. More importantly, we show here that iGH correlates to disease as well as if not better than PPD and thus we suggest that it could be a standalone metric of periodontal health. Of course, the true value will need to be validated with larger cohorts including molars that could not be accessed here due to the large size of the transducer.

Lastly, iGT was compared to gingival biotype for the patient set. Gingival thickness alone does not reflect periodontal health but is an important metric in the context of gum grafts and periodontal flap surgeries. Currently, biotype is a binary evaluation performed by inserting the periodontal probe into the gingival sulcus and assessing probe visibility. A visible probe corresponds to a “thin” biotype and an invisible probe corresponds to a “thick” biotype. Clinical consensus associates a thin biotype with <1.0 mm GT and a thick biotype with >1.0 mm GT. In our patient set, 93.5% of the associated gingiva for measured teeth possessed a thick biotype, and there was no correlation to disease status (**Fig. S2A**). Accordingly, the iGT measurements (taken from the midpoint of the ABC and GM) were not significantly different for healthy versus diseased patients (**Fig. S2B**). Though imaging is much more precise than the probe visibility method, this comparison served as assurance that iGT measurements were unbiased by the health status of the patient.

In our dataset, many images possessed reflection artifacts that are not representative of typical US images collected with a transducer sleeve. The reason for these artifacts was the specific geometry of the transducer, i.e., the ∼ 0.5-mm gap between the transducer elements and the subject/imaging target. Most transducers do not have this gap. Another limitation of the transducer was its size. This restricted imaging to the buccal surfaces of teeth 6-11 and 22-27. The ideal transducer could access the buccal and lingual surfaces of the full dentition. For future clinical deployment, it would also be desirable to integrate computational techniques for the automatic assessment of image quality and assignment of anatomical markers ^11, 17^. Despite these limitations in the current study, this technique may have significant clinical value for longitudinal monitoring of periodontal health. Unlike other oral imaging modalities, ultrasonography has the unique advantage of integrating signal from both hard and soft tissues, facilitating the measurement of periodontal metrics that require the resolution of both hard (ABC, CEJ) and soft (GM, GT) features. Further, it is non-ionizing, painless, and can be operated chairside with minimal training.

## CONCLUSIONS

We investigated the use of high-frequency US in 10 healthy subjects (34 teeth) and 6 subjects with periodontal disease (32 teeth) for measuring critical metrics of periodontal health, including probing pocket depth, clinical attachment level, gingival recession, and gingival thickness at midbuccal sites. Image-based measurements of gingival height extended from the gingival margin to the alveolar bone crest and were comparable to probing pocket depth (1.57-mm magnitude bias) with functional equivalence for assessing disease status. Identification of the cementoenamel junction by human operators also allowed image-based measurement of alveolar bone level and gingival recession. Interoperator bias was negligible (<0.1 mm) for gingival height and 0.45 mm for alveolar bone level measurements. Image-based alveolar bone level measurements were equivalent to clinical attachment level for staging disease (0.25-mm magnitude bias). Overall, ultrasonographic metrics in this patient group had at least an equivalent diagnostic capacity as gold-standard physical probing while offering more detailed anatomical information and painless operation. We anticipate that advances in the form factor of US hardware will facilitate the further translation of this technology into the dental clinic.

## Supporting information

The supporting information contains Supplemental Table 1 and Supplemental Figures 1 - 2.

## Data Availability

All data produced in the present study are available upon reasonable request to the authors.

## ACKNOWLEDGEMENTS

The authors acknowledge NIH funding under R21 DE029025, R21 DE029917, and UL TR001442. This publication was supported in part by the National Science Foundation Graduate Research Fellowship Program under Grant No. DGE-1650112. C.M. graciously acknowledges support from the ARCS Foundation. Panels A,C, and D of Figure 1 were created in part with BioRender.

