## Supplementary material for "High-resolution ultrasonography of gingival biomarkers for periodontal diagnosis in healthy and diseased subjects": The supporting information contains Supplemental Table 1 and Supplemental Figures 1 - 2.

**Table of Contents.**

**Table S1**. Image quality metrics and clinical measurements.

**Figure S1**. Comparison of ultrasound to physical examination for an extracted swine mandible following gum flap resection and periodontal measurement with physical probing.

**Figure S2**. Comparison between clinically assigned biotype and US image-based measurements of gingival thickness (iGT).

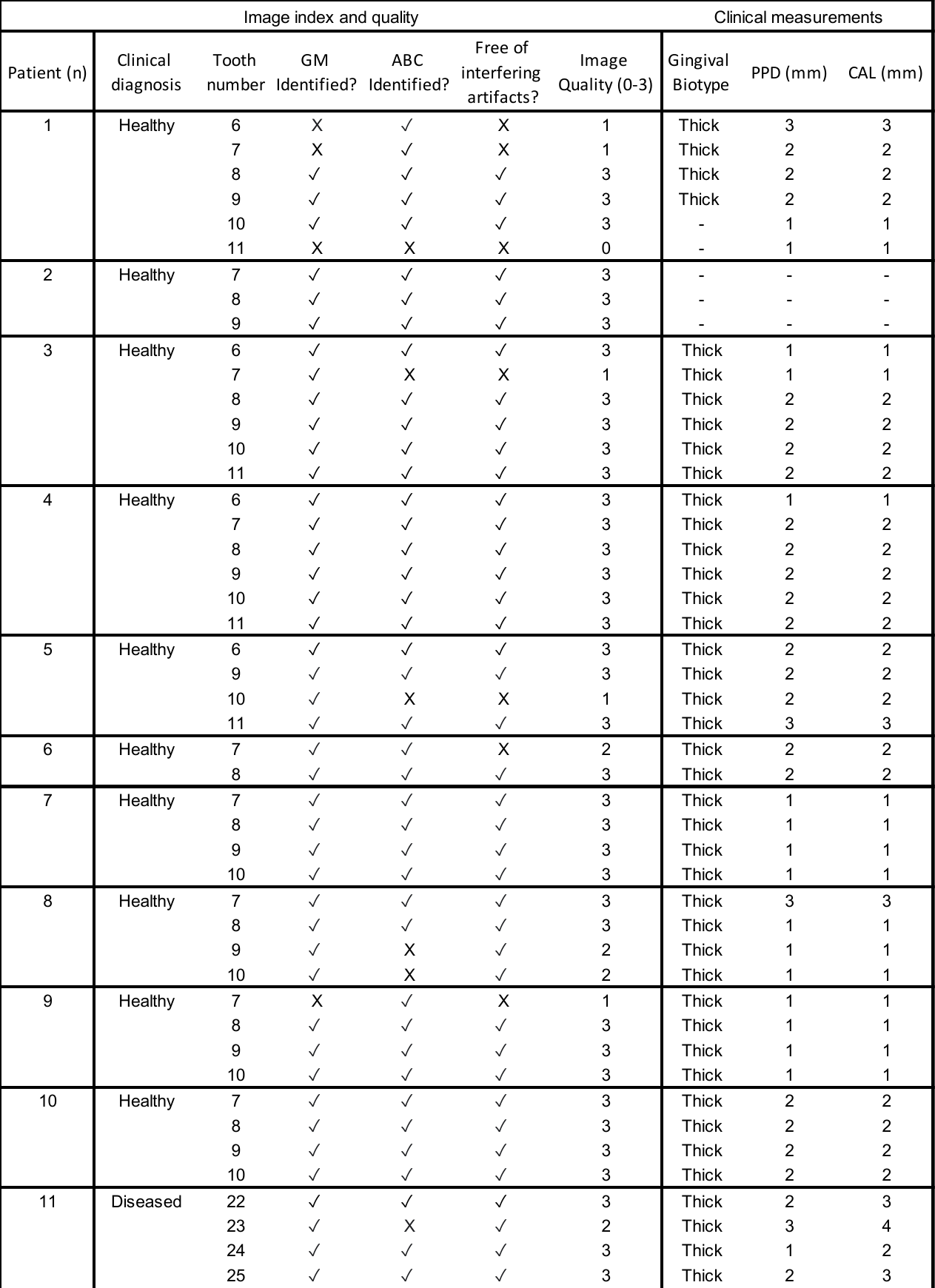

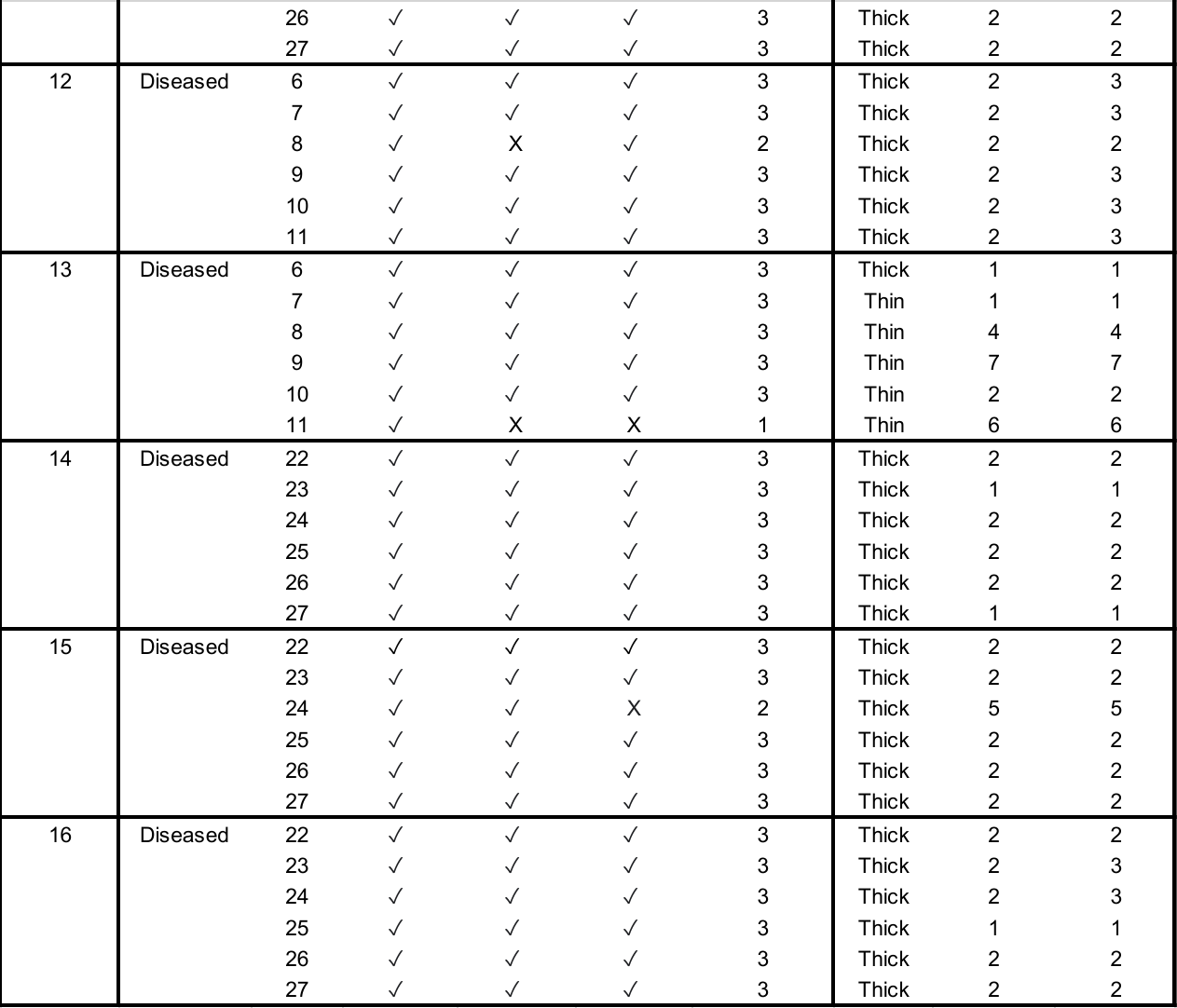

**Table S1**. Image quality metrics and clinical measurements.

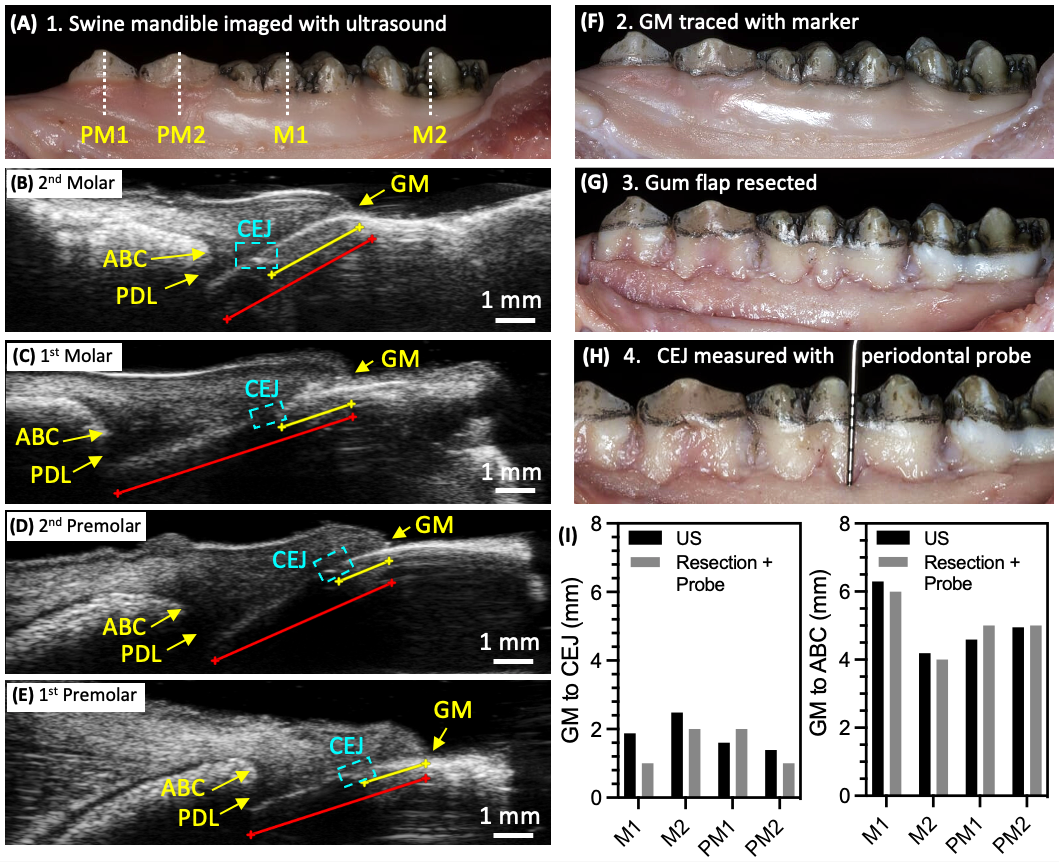

**Figure. S1. Comparison of ultrasound to physical examination for an extracted swine mandible following gum flap resection and periodontal measurement with physical probing.** (**A**) Photograph of the 2^nd^ molar (M2), 1^st^ molar (M1), 2^nd^ premolar (PM2), 1^st^ premolar (PM1), and the imaging plane for each tooth (white dashed lines). (**B-E**) Visualization of the cementoenamel junction (CEJ) relative to the alveolar bone crest (ABC) and gingival margin (GM) for the (**B**) 2^nd^ molar, (**C**) 1^st^ molar, (**D**) 2^nd^ premolar, and (**E**) first premolar. The CEJ is consistently resolvable as a disruption in the echogenicity of the tooth surface between the GM and the ABC. The image-based measurements of the GM to CEJ (yellow lines) were M2 = 2.53, M1 =1.88, PM2 = 1.39 mm, PM1 = 1.60 mm. The corresponding GM to AC measurements (red lines) were M2 = 4.19 mm, M1 = 6.30 mm, PM2 = 4.95 mm, PM1 = 4.59 mm. (**F**) Following imaging, the GM was traced along the teeth with a marker. (**G**) The gingiva (gum flap) was resected to reveal the roots of the teeth. (**H**) The distances from GM to CEJ and GM to ABC were measured by a clinician using physical probing. (**I**) These values were plotted and compared to the image-based measurements. All resection + probe values were restricted to integers.

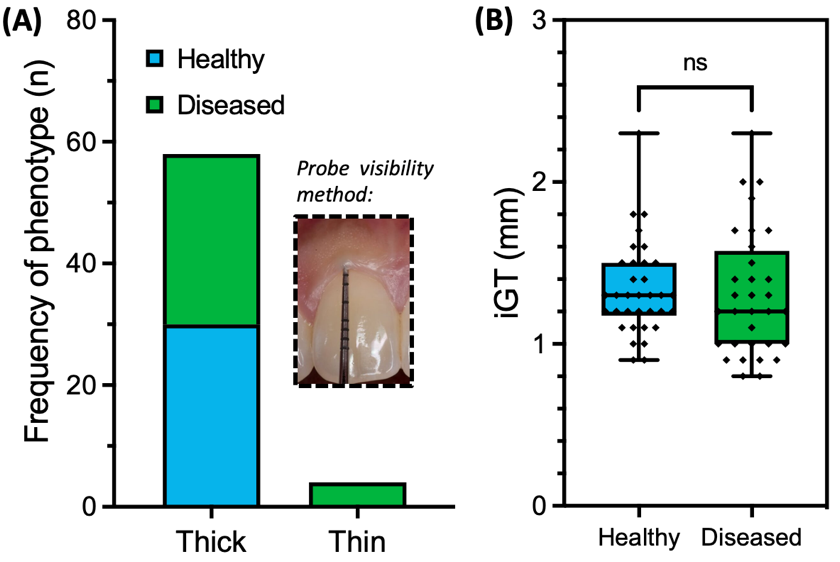

**Figure S2. Comparison between clinically assigned biotype and US image-based measurements of gingival thickness (iGT).** (**A**) Teeth from healthy and diseased groups were classified as a thick or thin biotype according to the conventional probe-visibility method. Photographic inset: example of a thick biotype. (**B**) The imaged gingival thickness (iGT) can provide significantly more quantitative assessment (< 0.1 mm precision over the full cross-sectional area) than probe-based biotyping. As expected, no significant difference was observed between iGT for the healthy and diseased groups. Here, the iGT was measured from the midpoint between the ABC and GM.
